# During outbreak periods, fever rates are lower in the morning, raising concerns about only conducting fever screenings at that time

**DOI:** 10.1101/2020.05.23.20093484

**Authors:** Charles Harding, Francesco Pompei, Samantha F Bordonaro, Daniel C McGillicuddy, Dmitriy Burmistrov, Leon D Sanchez

**Author notes:** **Corresponding Author:** Leon D Sanchez, MD, MPH; Beth Israel Deaconess Medical Center Emergency Department and Harvard Medical School; Boston, MA, USA.

## Abstract

Body temperatures are generally lower during mornings, but it is unclear how this affects practice during disease outbreaks. We retrospectively studied fever rates during seasonal influenza outbreaks and the 2009 H1N1 (swine flu) pandemic, analyzing Boston emergency department visits (2009– 2012; *n*=93,225) and a nationally representative sample of adult US emergency department visits (National Hospital Ambulatory Care Survey, 2002–2010; *n*=202,181). Outbreak periods were defined from regional and national ILINet thresholds set by the Centers for Disease Control and Prevention. During outbreak periods, temperatures were about half as likely to reach the fever range (≥100.4°F, ≥38.0°C) in the morning as in the evening (rate ratios for 6 AM–noon vs. 6 PM– midnight: Boston=0.43, 95% CI=0.29-0.61; national=0.56, 95% CI=0.47-0.66; national with multivariable adjustment for 12 case characteristics=0.59, 95% CI=0.50-0.70). Fever rates were also lowest during mornings for other common fever definitions and in supplementary analyses of a non-emergency-department, non-medical population of adults (National Health and Nutrition Examination Survey I). Our results suggest that mornings may be a bad time to perform once-daily fever screenings for infectious diseases, and that twice-daily screenings could be preferable as a simple solution. However, similar research is needed on COVID-19 to address the current pandemic.

**One Sentence Summary:** Fevers are about half as common in the morning as in the evening during influenza outbreaks, suggesting that mornings may be a bad time to perform once-daily fever screenings for infectious diseases, and that twice-daily screenings could be preferable.

## Introduction

Body temperatures are less likely to reach the fever range during mornings^1–5^, but it is unknown how this affects fever detection during disease outbreaks. We retrospectively investigated fevers during seasonal influenza outbreaks and the 2009 H1N1 (swine flu) pandemic, which have been used as preparatory models for coronavirus disease 2019 (COVID-19). We sought to compare fever rates by time of day during outbreak periods in records from a Boston emergency department^2^ (September 2009 to March 2012) and a nationally representative sample of adult US emergency department visits (December 2002 to December 2010)^6^. This brief report builds on a study of the same data sources that predates COVID-19 and lacks analyses of time-of-day fever changes during outbreaks^2^.

## Results

In total, 93,225 and 202,181 temperatures were analyzed from the Boston and national studies, respectively. At the Boston and national emergency departments, 54% and 54% of patients were women, and the median patient ages (inter-quartile range) were 49 (32-66) and 34 (18-52) years, respectively (evaluation periods: Boston, September 2009 to August 2011; national, December 2002 to December 2010). More detailed patient characteristics are given in our earlier study^2^.

In all investigated periods, fever-range temperatures (≥100.4°F, ≥38.0°C) were less common during mornings than during evenings (**Fig. 1; Supplementary Table 1**). Morning-evening changes were especially large during influenza outbreaks (ratio of 6 AM–noon vs. 6 PM–midnight: Boston=0.43, 95% CI=0.29-0.61; national=0.56, 95% CI=0.47-0.66). Because temperatures were taken from different patients at different times of day, we used multivariable logistic regressions to adjust for time-of-day changes in 12 case mix characteristics when analyzing the national data. Results were not substantially changed (adjusted morning-evening ratio=0.59, 95% CI=0.50-0.70). Findings were also similar when analyzing other fever definitions used for COVID-19 (**Fig. 2**).

**Figure 1.**
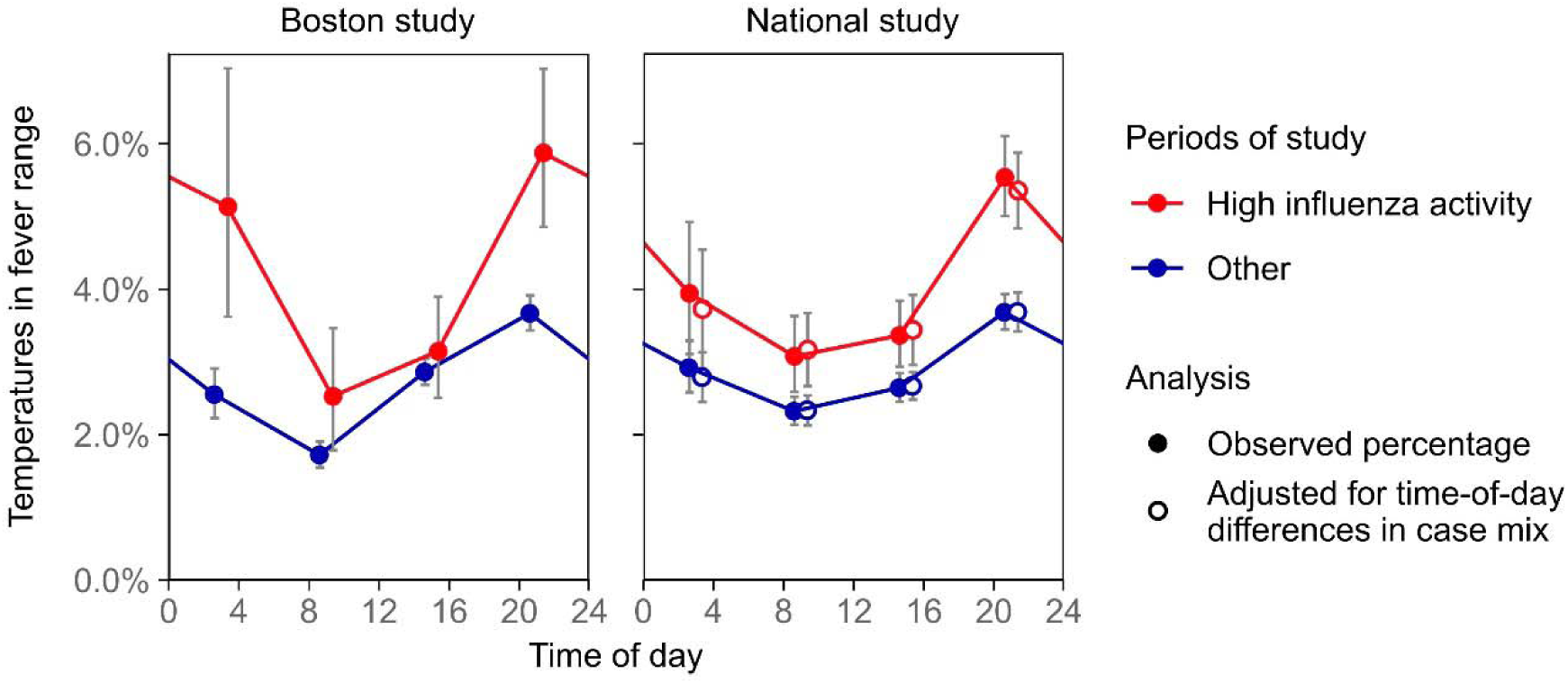
Time-of-day changes in the percentage of body temperatures that reach the fever range: Boston and US national studies. In both the Boston and US national studies, temperatures measured during mornings were less likely to reach the fever range (≥100.4°F, ≥38.0°C), especially during periods of high influenza activity (seasonal flu and the 2009 H1N1 pandemic). Overall, temperatures were roughly half as likely to meet the definition of fever in the morning as in the evening (**Supplementary Table S1**). The results suggest that morning temperature measurements could miss many febrile disease cases, which raises concerns because workplace and school fever screens often occur during mornings, and because patients seen for potential COVID-19 may only have temperatures checked during mornings. A simple solution is twice-daily temperature measurement. National study results are nationally representative of adult visits to US emergency departments. Confidence intervals are 95%. Overlapping points were shifted slightly on the x axis for clarity.

**Figure 2.**
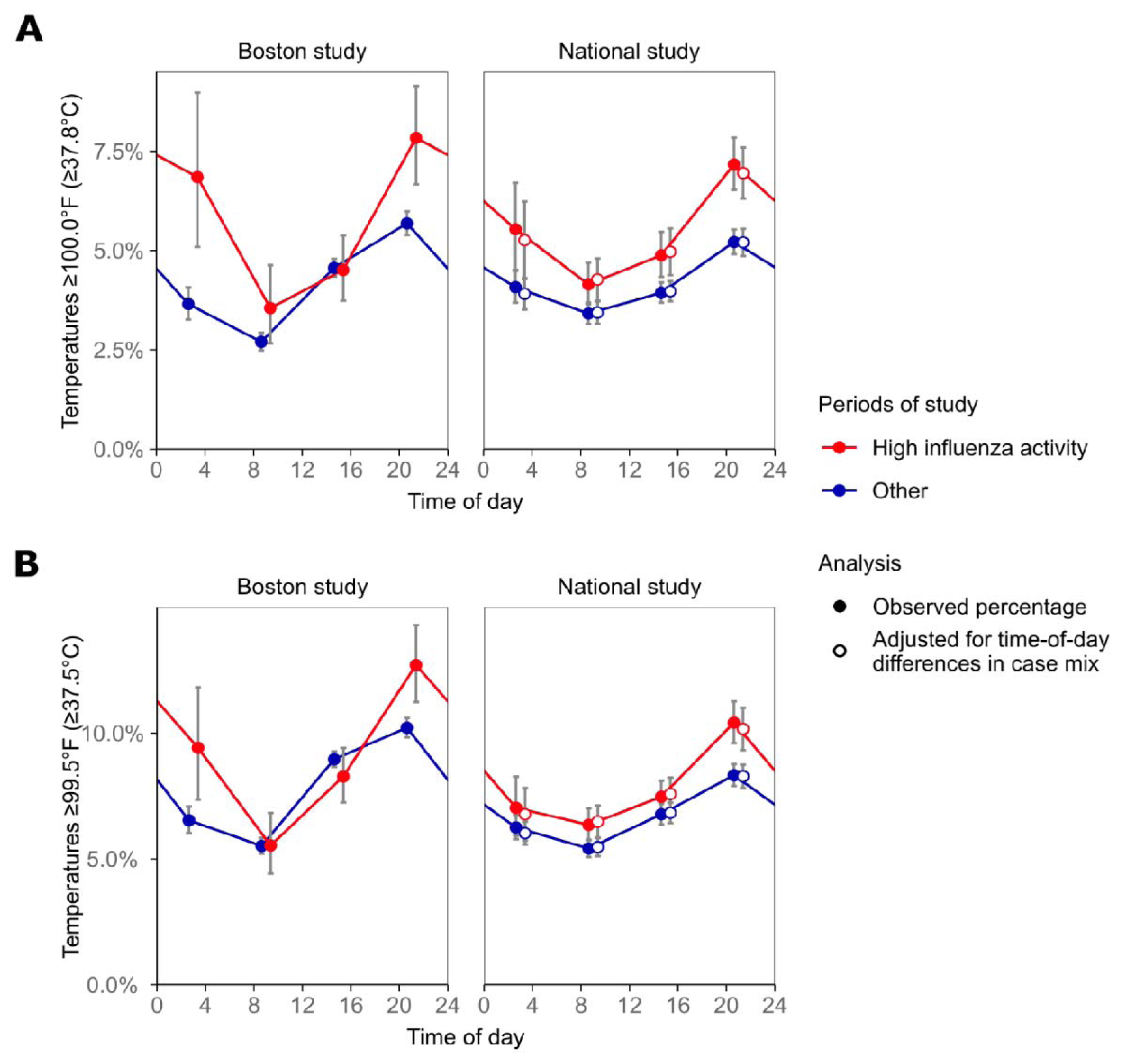
Time-of-day changes in fever rates using other fever definitions that are commonly applied for COVID-19. (A) When the fever definition is lowered to ≥100.0°F (≥37.8°C), large time-of-day changes in fever rates are still observed, especially during periods of high influenza activity (influenza-period ratio of fever rates at 6 AM–noon vs. 6 PM–midnight: Boston=0.45, 95% CI=0.32-0.61; national=0.58, 95% CI=0.50-0.67; case-mix-adjusted national=0.61, 95% CI=0.53-0.71). (B) When the fever definition is further lowered to ≥99.5°F (≥37.5°C), time-of-day changes in fever rates continue to be observed, including during high influenza activity (influenza-period ratio of fever rates at 6 AM–noon vs. 6 PM–midnight: Boston=0.44, 95% CI=0.34-0.55; national=0.61, 95% CI=0.54-0.69; case-mix-adjusted national=0.64, 95% CI=0.56-0.72). However, more cases are classified as having fever and fever rates during high influenza activity are no longer as distinguishable from fever rates during other periods. This may be because the lower threshold includes more individuals who do not physiologically have fever (false positives). If false positives are too common, they can be an obstacle to screening. Confidence intervals are 95%. Overlapping points were shifted slightly on the x axis for clarity.

Supplementary investigations showed similar results when analyzing time as a continuous variable (**Supplementary Fig. 1**) and evaluating years separately (**Supplementary Fig. 2**). Additionally, investigations of Berkson’s bias (collider selection bias) and residual confounding supported the main findings, including in analyses of an additional data source with body temperatures from the general (non-medical) US population (*n*=6535; **Appendix, Berkson’s Bias and Residual Confounding**).

## Discussion

Our results raise concerns that morning measurements could miss many (perhaps even half) of the individuals with fevers detectable during evenings, potentially allowing them to go to work, attend school, and travel. Physiologically, circadian rhythms usually reach temperature low points during mornings, and patients can lack fever signs or can present some signs without reaching cutoffs like ≥100.4°F (≥38.0°C)^1–5,7,8^. Although circadian rhythms are well known, their relevance to fever is less recognized because it was less important before COVID-19.

Temperature screenings are used for COVID-19 because measurements are simple, fever is common and presents early^9,10^, and many symptomatic people do not self-isolate^11,12^. A summary of COVID-19 fevers is provided in the appendices (**Appendix, COVID-19 Fevers**). Temperature and other symptom screenings are limited by an inability to detect presymptomatic or asymptomatic cases. However, especially when layered together, partially effective measures can confer meaningful benefits by reducing COVID-19 transmission rates—a justification that also underpins public use of face masks, 6-foot physical distancing, and handwashing (**Appendix, Screening and Transmission)**. Additionally, the potential usefulness of symptom screening can be heightened in healthcare settings, for example as evidenced by a report that about two-thirds of Seattle-area healthcare personnel with symptomatic COVID-19 kept working after developing symptoms (work duration with symptoms: median, 2 days; range, 1-10 days)^13^.

Temperature screening is usually recommended once daily at morning arrival to workplaces and schools, yet our results suggest the morning could be the worst time. A rapidly applicable solution may be twice-daily screening, for example before and after shifts. The first measurement is retained to reduce possible during-shift transmission, and the second is for cases previously missed. With two widely spaced measurements, at least one avoids the temperature low point, regardless of individual differences in shift and circadian rhythm timing. (Both can be large in some groups, such as night workers.) An alternative solution could be once-daily screening with a revised definition of fever that is lower in the morning. However, lowering the morning fever definition is an averaged, population-level correction that does not address individual differences in circadian timing. Additionally, the only available morning-lowered fever definition^1^ appears to overcorrect^2^. Irrespective of the chosen solution, self-measurements and symptom checks at home may help meet privacy regulations and reduce burdens at workplaces and schools.

Given the possibility of missing fevers during mornings, evening temperature remeasurements might be requested at morning COVID-19 examinations, an approach that could be useful where SARS-CoV-2 testing is limited to febrile patients because of shortages. Similarly, departure and arrival screens might both be worthwhile for long flights—an option suggested previously to address symptom changes during flight^14^.

Our findings should be interpreted with several cautions: First, our results are from clinicians using hospital-grade thermometers, and may not generalize to other settings, layperson measurements, or low-accuracy, non-contact thermometer guns and thermal imagers. Second, screenings should balance false-negative risks with false-positive burdens, which could increase during evenings when healthy temperatures rise^1,2^. Third, our analyses may not fully address confounding and Berkson’s biases, which could contribute artifactually to morning-evening fever differences. A relevant improvement could be to study temperatures longitudinally in the same patients, but inpatients are the only population with temperature measurements throughout day and night, and their circadian rhythms are often severely disrupted by hospital environments^15^. Fourth, thermometer site, age, and other factors also affect temperature^7,16^. Screenings may benefit from adjustment for some of these factors, especially thermometer site. Lastly and most importantly, although most diseases include morning temperature lows, this has not been shown for COVID-19. We hope our research encourages study of COVID-19 fevers and optimal screening strategies, especially to help workplaces and schools stay open where COVID-19 has been regionally controlled, and to help limit disease transmission in healthcare settings.

## Methods

### Data and definitions

Temperatures (*n*=115,149) were recorded during triages to monitor outbreaks at a Boston adult emergency department (September 2009–March 2012)^2,17^. We also investigated adult triage temperatures (*n*=218,574) from a nationally representative study of US emergency department visits (December 2002–December 2010)^6^. The thermometer types used were contact-type temporal artery (Boston) and a nationally representative sample (national). We analyzed exact measurement times (Boston) or arrival times as a substitute (national). We excluded records missing temperature or time (Boston=1.0%, national=7.5%), or indicating repeated or accidental measurement (repeated ≤15 seconds or temperature <95°F: Boston=18.0%), leaving 93,225 Boston and 202,181 national temperatures for analysis^2^. High-influenza activity periods were defined as months fully exceeding the ILINet baseline thresholds set by the Centers for Disease Control and Prevention for region 1 (Boston analysis; outbreak-period *n*=6627) or nationally (national analysis; outbreak-period *n*=29,908).^18^ This brief report extends a study that predates COVID-19 and analyzed daily fever cycles in the same datasets, but did not examine outbreak periods^2^. The earlier study also lacks our supplemental analyses of continuous time, separate years, and general (non-medical) population body temperatures. However, the earlier study includes additional methodological detail, patient demographics, and analyses demonstrating robustness to exclusion criteria choices.

### Ethics declarations

The institutional review board of Beth Israel Deaconess Medical Center approved the Boston study with a waver of informed consent. National analyses are of publicly available de-identified data. The research complies with all relevant ethical regulations.

### Statistical analysis

Nationally representative results were obtained by accounting for the national study’s multistage design^6,19^. Using multivariable logistic regression^19,20^, time-of-day case mix differences in 12 characteristics were excluded from responsibility for the time-of-day fever rate differences in national data (characteristics: age, case urgency, pain, sex, race, Hispanic or Latino ancestry, hospital admission, test ordering, procedure administration, medication ordering, ambulance arrival, and expected payment source; **Appendix Methods**). Anonymity requirements prevented multivariable analyses of Boston data.

### Data availability

Anonymized datasets from the National Hospital Ambulatory Medical Care Surveys (NHAMCS) and National Health and Nutrition Examination Survey I (NHANES I) are publicly available from the National Center for Health Statistics of the U.S. Centers for Disease Control and Prevention (www.cdc.gov/nchs/ahcd/datasets_documentation_related.htm; wwn.cdc.gov/nchs/nhanes/nhanes1/default.aspx). Data from the Boston study may be available from the authors upon reasonable request, subject to privacy restrictions.

## Supporting information

STROBE checklist

## Data Availability

Anonymized datasets from the National Hospital Ambulatory Medical Care Surveys (NHAMCS) and National Health and Nutrition Examination Survey I (NHANES I) are publicly available from the National Center for Health Statistics of the U.S. Centers for Disease Control and Prevention. Data from the Boston study may be available from the authors upon reasonable request, subject to privacy restrictions.

https://www.cdc.gov/nchs/ahcd/datasets_documentation_related.htm

https://wwwn.cdc.gov/nchs/nhanes/nhanes1/default.aspx

## Author contributions

C.H., F.P., D.B., and L.D.S. contributed to conception and design of the study; S.F.B., D.C.M., and L.S. were involved in data collection; C.H. and D.B. were involved in data analysis; C.H. drafted the article; C.H., F.P., and L.D.S. were involved in critical revisions. All authors reviewed the manuscript.

## Competing interests

Exergen, Corp. (Watertown, MA) provided thermometers (temporal artery, model TAT-5000), technical support and funding, including funding for C.H. and D.B.’s participation. F.P. is CEO and founder of Exergen and holds patents related to the devices under study. C.H. and D.B. have been consultants to Exergen. Exergen was involved in conceptualization, design, data collection, analyses, drafting, and publication decisions. There are no other funding sources or conflicts of interest to disclose for this report or the originating studies.

## Notes

### Author Declarations

The institutional review board of Beth Israel Deaconess Medical Center approved the Boston emergency department study (2008-P-000412).

### Summary of Updates

Abstract expanded; supplemental appendices updated to include newer references; article retitled; article reformatted per journal requirements.

