## Supplementary for "During outbreak periods, fever rates are lower in the morning, raising concerns about only conducting fever screenings at that time"

**Contents** **Page**

Supplementary Table S1. Morning-evening comparisons........ 2
Supplementary Figure S1. Continuous time analysis........ 3
Supplementary Figure S2. Year-by-year analysis........ 4
Appendix, Berkson's Bias and Residual Confounding........ 6
Figure A. Weekdays vs. weekends analysis........ 9
Figure B. Fever rates in the general population analysis........ 13
Appendix, COVID-19 Fevers........ 16
Appendix, Screening and Transmission........ 21
Appendix Methods........ 24
References........ 27

**Supplementary Table S1.** Fever rates in the Boston and national studies, showing consistent increases from mornings (6 AM to noon) to evenings (6 PM to midnight), with risk ratios for the morning-evening comparison ranging from 0.43 to 0.66. When evaluated on an absolute scale, morning-evening increases were largest during high influenza activity.

| Fever definition | Period | Boston study | | | National study, observed | | | National study, case-mix adjusted | | |
|---|---|---|---|---|---|---|---|---|---|---|
| | | Morning | Evening | RR (95% CI) | Morning | Evening | RR (95% CI) | Morning | Evening | RR (95% CI) |
| ≥100.4°F, ≥38.0°C | High influenza | 2.5% | 5.9% | 0.43 (0.29-0.61) | 3.1% | 5.5% | 0.56 (0.47-0.66) | 3.2% | 5.4% | 0.59 (0.50-0.70) |
| | Other | 1.7% | 3.7% | 0.47 (0.42-0.53) | 2.3% | 3.7% | 0.63 (0.57-0.69) | 2.3% | 3.7% | 0.63 (0.57-0.70) |
| ≥100.0°F, ≥37.8°C | High influenza | 3.6% | 7.8% | 0.45 (0.32-0.61) | 4.1% | 7.2% | 0.58 (0.50-0.67) | 4.2% | 7.0% | 0.61 (0.53-0.71) |
| | Other | 2.7% | 5.7% | 0.48 (0.43-0.52) | 3.4% | 5.2% | 0.65 (0.60-0.71) | 3.4% | 5.2% | 0.66 (0.60-0.72) |
| ≥99.5°F, ≥37.5°C | High influenza | 5.5% | 12.7% | 0.44 (0.34-0.55) | 6.4% | 10.4% | 0.61 (0.54-0.69) | 6.5% | 10.2% | 0.64 (0.56-0.72) |
| | Other | 5.5% | 10.2% | 0.54 (0.50-0.58) | 5.4% | 8.3% | 0.65 (0.61-0.70) | 5.5% | 8.3% | 0.66 (0.62-0.71) |

RR, risk ratio comparing mornings to evenings. CI, confidence interval.

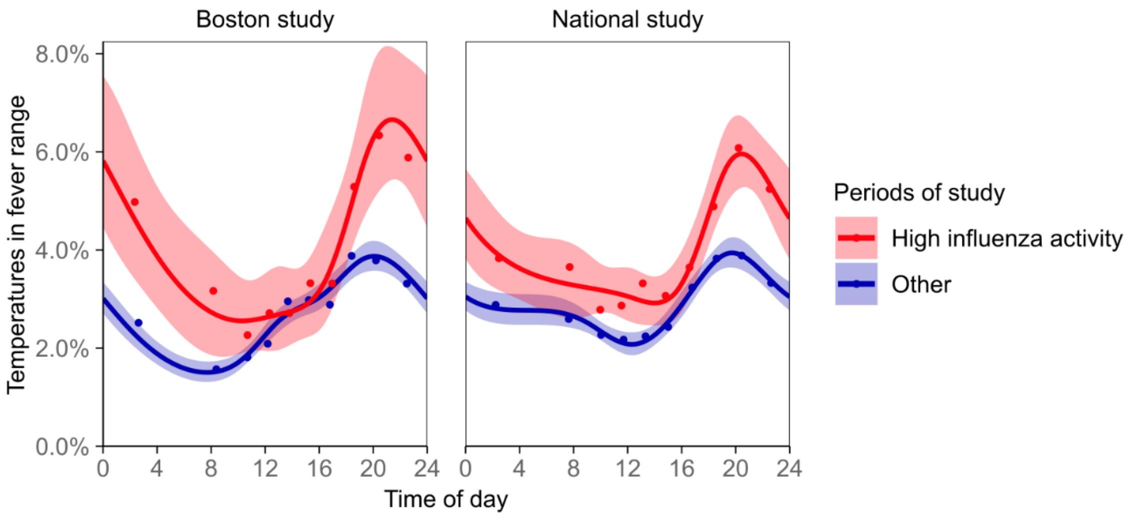


**Supplementary Figure S1. Time-of-day variation in the rate of fever (temperature ≥100.4°F, ≥38.0°C), with time analyzed as a continuous variable.** Results are similar to the binned analysis in the main paper (Figure 1), but show the cycle of fever rates over the day with more detail. Curves are from logistic regressions using a cyclic cubic spline term with knots placed at quintiles of the recorded times of day and midnight. To illustrate the correspondence between the data and the curves, points are also shown with the average time and fever rate for every 10% segment of the recorded times of day. As in the previous figures, national study results are nationally representative of adult visits to US emergency departments. Confidence bands are 95% (pointwise).

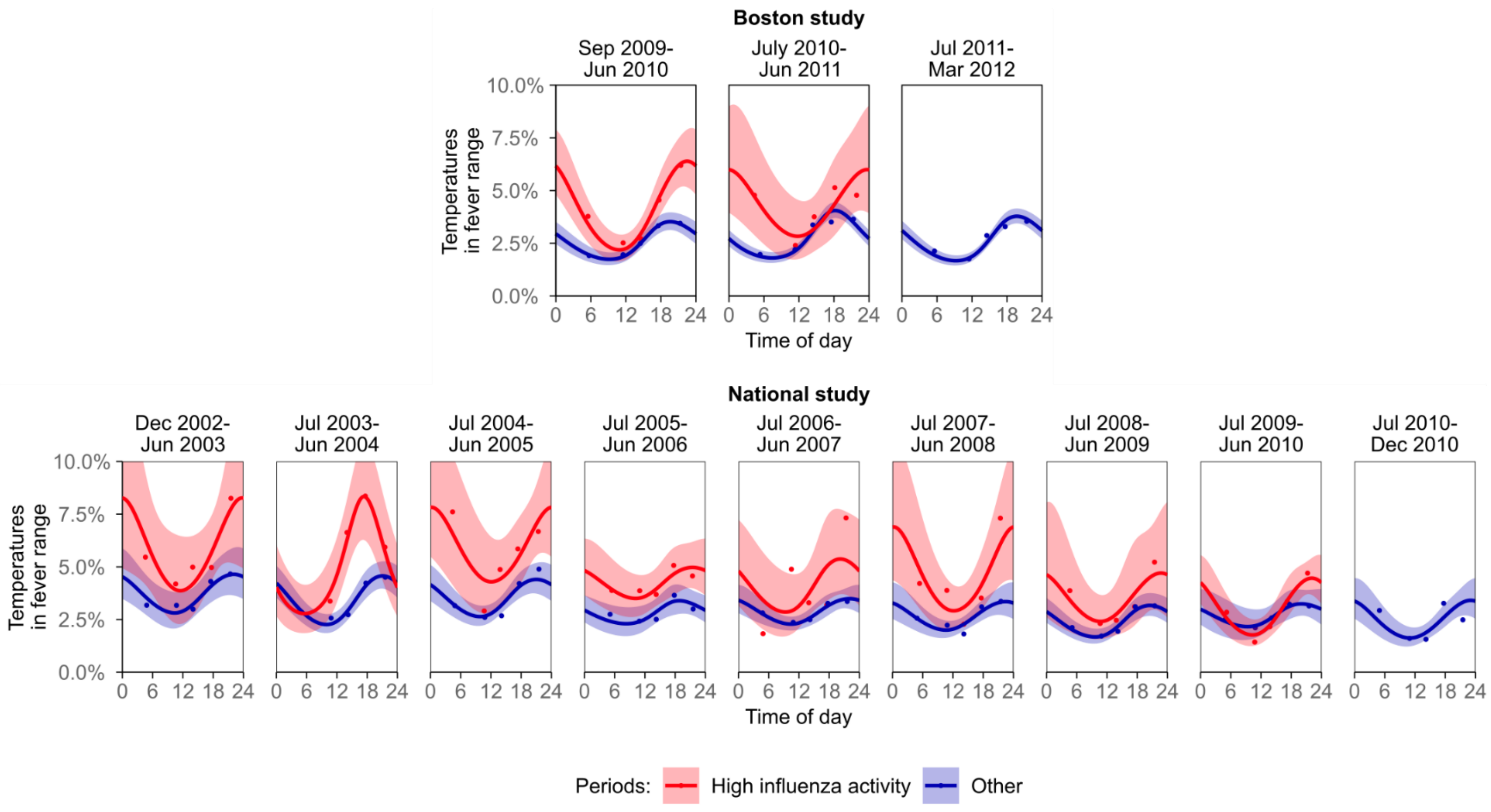


**Supplementary Figure S2. Time-of-day variation in the rate of fever (temperature ≥100.4°F, ≥38.0°C) during each year of the Boston and National studies.** In each year, morning temperatures were less likely to reach the fever range than evening temperatures, with especially large morning-evening differences usually occurring during high influenza activity. Fever rates in non-influenza periods were consistent across years of the studies. However, there were substantial year-to-year differences in overall fever rates during high influenza activity, which may be due to both year-to-year differences in the extent of influenza's spread and differences between each influenza virus's ability to cause fever. The high influenza activity period of 2009-2010 includes the influenza A(H1N1) pdm09 (swine flu) pandemic. In the figure, years are counted from July of one year through June of the next in order to keep temperatures recorded during the same flu seasons together. (Flu seasons occur in winter, so would be mixed across multiple plots if standard calendar years had been used instead.) In some years, no months of the study periods met our definition of high influenza activity due to unusually mild flu seasons (occurring in 2011-2012) or due to a flu season beginning too late (occurring in 2010-2010 of the national data). Curves are from logistic regressions using a cyclic cubic spline term with knots placed at tertiles of time of day and midnight. To illustrate correspondence between the data and the curves, points are also shown with the average time and fever rate for every 20% segment of the recorded times of day. The Boston study lasted from September 2009 to March 2012, while the national study lasted from December 2002 to December 2010. All confidence bands are 95% (pointwise).

### Appendix, Berkson's Bias and Residual Confounding

This appendix addresses topics of Berkson's bias and residual confounding. To do so, it includes analyses of weekday vs. weekend fever rates and fever rates in a large U.S. study of the general (non-medical) population ($n$=6,535), which may be of independent interest to some readers.

*Background*

In this study, the analyzed body temperatures were taken from patients presenting to emergency departments. Consequently, the morning and evening temperatures come from different groups of patients—namely, from patients who went to emergency departments in the morning and from those who went to emergency departments in the evening. This raises the possibility that the differences between the observed morning and evening fever rates do not result from differences in the actual rates of fever at these times, but instead result from other differences between the patients seen during mornings and evenings.

In the main text, we used a multivariable logistic regression approach to control for time-of-day differences in 12 variables. This controlling produced almost no change in the fever rates, which substantially raises our confidence that the findings do not result from time-of-day differences in the case mix of patients seen at emergency departments. However, it remains possible that the controlling was not sufficient to address confounding or that the findings are affected by selection bias in the form of Berkson's bias—possibilities addressed in this appendix.

There are several names for Berkson's bias, including Berkson's fallacy, Berkson's paradox, Berksonian bias, collider selection bias, and collider stratification bias.[1,2] Berkson's bias may alter the estimated relationship between an exposure and outcome when a study is limited to a patient group, and when the exposure and outcome affect membership in that group.[2] For the current study, the bias could apply because the study is limited to patients presenting to emergency departments (patient group), and because both time of day (exposure) and body temperature (outcome) can affect the decision to go to an emergency department. Explained in common language, Berkson's bias could be expected to arise in the following way: If it is harder to go to the emergency department at some times of day than others, a more substantial collection of symptoms may be necessary to induce an emergency department visit at these times. Since the presence of fever may make an individual's overall extent of symptoms more substantial, this could result in an inverse association between convenient times of day and fever rates, unrelated to the rhythms of body temperature in disease.

For technical discussion of Berkson's bias, we refer the reader to articles by Westreich[2] and Snoep et al.[1] As these show, an important feature of Berkson's bias is that it is not eliminated by using multivariable regression to control for confounders. Instead, it can be addressed by considering the underlying logic of the situation, through sensitivity analyses, and by analyzing a general population (nonmedical) dataset, and by referring to previous research. These points are also relevant to residual confounding. We address them below.

*Underlying logic*

In our study, fever rates are higher at the more convenient times of day (after work and during the evening; **Figure 1, Supplementary Figure S1**). This is contrary to the effect we generally expect from Berkson's bias, but is consistent with the circadian rhythm of body temperature.[3] Considering the underlying logic in this way suggests that, if present, Berkson's bias would be likely to dampen the size of the morning-evening difference, rather than inflate it.

*Sensitivity analyses*

We compared fever rates during weekdays and weekends as a check on the potential effects of Berkson's bias and, more generally, the effects of differences between weekday and weekend schedules on the observed fever rates. Such effects could arise, for example, since the decision to go to the emergency department can be affected by work and schooling hours, and by the hours and days that alternative sources of care are open (such as primary care physicians' offices). Work shifts, school times, and alternative care availability are the main mechanisms through which we anticipate Berkson's bias and residual confounding could occur during daytime hours, and we therefore think the weekday vs. weekend comparison should be a revealing assessment for both.

We found that the time of day variation in fever rates was similar on weekdays and weekends, as shown in Figure A below. The similarity between weekday and weekend results is contrary to the anticipated mechanisms of Berkson's bias and residual confounding, but matches with the physiological consistency that is expected for the circadian rhythm of body temperature.

Our previous research also includes comparisons of weekend and weekday fever rates for other fever definitions, also with similar results.[3]

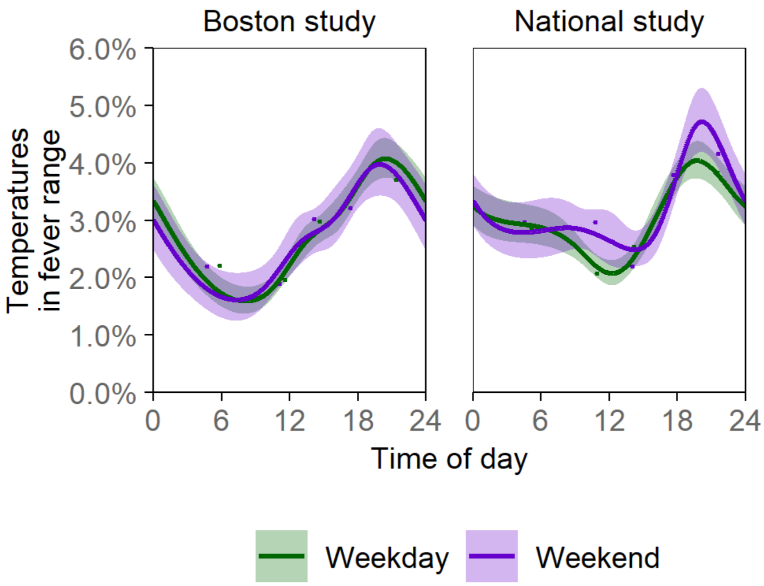


**Appendix on Berkson's bias, Figure A. Time-of-day variation in the rate of fever (temperature ≥100.4°F, ≥38.0°C), comparing weekdays and weekends.** The time-of-day cycle of fever rates is similar during weekdays and weekends, suggesting that the cycle of fever rates is not a consequence of daily schedule changes or the availability of alternative sources of care. Curves are from logistic regressions using a cyclic cubic spline term with knots placed at quintiles of the recorded times of day and midnight. To illustrate the correspondence between the data and the curves, points are also shown with the average time and fever rate for every 20% segment of the recorded times of day. As in the previous figures, national study results are nationally representative of adult visits to US emergency departments. Confidence bands are 95% (pointwise).

*Analysis of a general population (nonmedical) dataset*

As a further check for Berkson's bias, we analyzed body temperatures from a large general population (non-medical) study of the United States, the National Health and Nutrition Examination Survey (NHANES) I. Analyzing general population body temperatures is also useful to examine the generalizability of our emergency department findings.

*Methods:* NHANES I was performed in 1971-1975 to evaluate the health of the general US population. Despite the age of this study, it appears to be the largest collection of general population body temperatures that is currently available. Perhaps because of this, NHANES I body temperatures have been the subject of renewed interest, including in a prominent study by Protsiv et al.[4] Clinical examination data are available from 23,808 persons age ≤75. Time of day was recorded during clinician-performed blood pressure assessments that were given to 6,877 adults, all of whom were age 25-75. Of the adults with time of day records, 339 were missing body temperature measurements, 2 had unrealistically low body temperatures indicating measurement error (<95.0°F, <35.0°C), and 1 was missing information on obesity status. We excluded these individuals and analyzed body temperatures for the remaining 6,535 persons.

Analyses of NHANES I can provide nationally representative findings by accounting for the survey's design. However, because temperatures and times were only available for a subset of individuals, we chose not to account for the survey design in the analyses, meaning that the results are not nationally representative estimates, but should instead be interpreted as coming from a cohort study of 6,535 persons.

Body temperatures were measured by a clinician using oral mercury thermometers. Because of the non-medical population under study, high temperatures were much rarer than in the emergency department data. Of the 6,535 analyzed individuals, 1 had a temperature ≥100.4°F (≥38.0°C), 6 had a body temperature ≥100.0°F (≥37.8°C) and 30 had a body temperature ≥99.5°F (≥37.5°C). The lack of temperatures ≥100.4°F (≥38.0°C) prevented analyses of this fever threshold, and analyses were therefore restricted to ≥100.0°F (≥37.8°C) and ≥99.5°F (≥37.5°C).

The examinations were performed from morning to evening, with 95% of all times of day falling between 9:05 AM and 9:20 PM. Older individuals were somewhat less common at evening examinations, and multivariate adjustment was therefore performed, following the same approach used for the NHAMCS national study of emergency department data.[5] The following covariates were included in the multivariate adjustment: gender (coded as male or female in NHANES I), age (analyzed a continuous variable using a spline with knots at ages 35, 45, 55, and 65), race (coded as black, white, or other in NHANES I), and obesity (coded as present or absent in NHANES I).

*Results:* As shown in Figure B, fever rates increased from morning (before noon) to evening (after 6 PM) for both investigated fever definitions. Multivariate-adjusted analyses continued to show increased fever rates from the morning to the evening. For the ≥99.5°F (≥37.5°C) fever definition, the morning-to-evening fever risk ratio was 0.23 (95% CI 0.09-0.64) in the unadjusted analysis and 0.24 (95% CI 0.09-0.67 in the adjusted analysis). For the ≥100.0°F (≥37.8°C) fever definition, the morning-to-evening fever risk ratio was 0.00 (95% CI 0.00-0.86 ) in the unadjusted analysis and was statistically undefined in the adjusted analysis because no fevers in this range occurred during mornings.

*Discussion:* Analysis of body temperatures from the general population continued to show increased fever rates from morning to evening, providing further support for a physiological origin of this morning-evening difference, rather than the alternative that it results from Berkson's bias.

However, there are several limitations to the general population results, including the rarity of fever-range temperatures in the studied cohort and the lack of overnight temperatures. Additionally, it is not clear whether NHANES I fever rates accurately estimate fever rates in the general population, or if individuals with fever were predisposed to not show up to their NHANES I examinations because they were sick. Yet, even if the latter possibility is true, it would be expected to lead to a reverse of the effects of Berkson's bias for emergency departments (where fever increases the chance of presentation, rather than decreasing it), meaning that the observation of morning-evening fever rate increases in both the general population and emergency department studies continues to support a physiological origin to this pattern.

In summary, findings from a general population cohort are consistent with morning-evening increases in fever rates that are physiological, rather than an artifact of Berkson's bias. Results also show that the morning-evening increases in fever rates occur outside of medical populations.

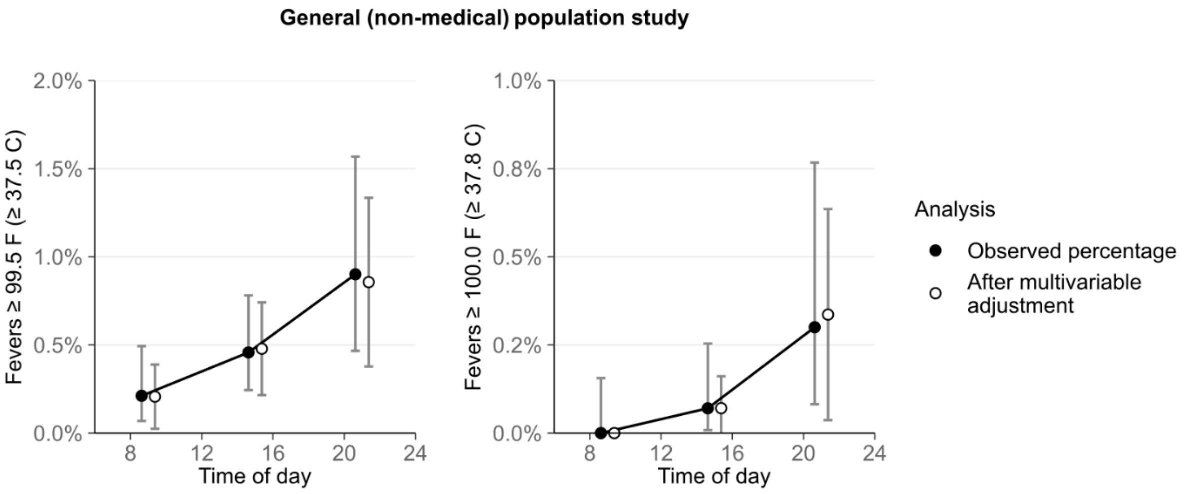


**Appendix on Berkson's bias, Figure B. Increasing fever rates by time of day in the general population study.** When analyzing a large (*n*=6,535) general (non-medical) population sample, the rate of fever-range temperatures continued to rise from morning to evening. This supports a physiological origin for the morning-evening difference in fever-range temperatures, rather than the alternative that Berkson's bias explains the morning-evening difference. However, there were too few fevers ≥100.4°F (≥38.0°C; *n*=1) to be analyzed by time of day in the general population study, and the general population study also did not include post-midnight temperature measurements. Confidence intervals are 95%. The multivariable analysis adjusts for age, sex, race, and obesity status, but does not differ meaningfully from the unadjusted results.

*Previous research*

In previous studies of the relationship between the circadian rhythm and fever, longitudinal data were collected on groups of hospital inpatients[6–8] and healthy young men with experimentally induced fevers.[9] Because of the longitudinal data collection in these studies, their results are not subject to Berkson's bias or confounding for morning-evening comparisons. Each study also showed a rise in fever-range temperatures from morning to evening, consistent with a physiological effect, though the sample sizes were not large enough to reliably evaluate the degree of morning-evening differences in those studies.

Drawing on previous research, numeric estimation also suggests that circadian physiology can cause fever-range temperatures (≥100.4°F, ≥38.0°C) to be half as common in the morning as in the evening—consistent with the morning-evening change observed in our study, without involving any Berkson's bias or residual confounding. The estimation proceeds as follows: Healthy body temperatures reportedly change an average of 0.9°F (0.5°C) from the morning low to the evening high as part of the physiology of the circadian rhythm.[10] Additionally, the morning-evening temperature change in febrile disease is usually at least as large or larger than observed in health.[11–14] In our Boston study, of all the body temperatures that were in the fever range in the evening (6 PM−midnight), 44% were at least 0.9°F above the fever threshold, and therefore would stay in the fever range in the morning if they reduced the average circadian amount. Similarly, in the national emergency department study, 48% of evening fever-range temperatures were at least 0.9°F above the fever threshold, and therefore would stay in the fever range in the morning if they reduced the average amount. So in both cases, estimation suggests that fever-range temperatures can be half as common in the morning as in the evening owing to circadian physiology alone, without any biases.

To examine the dependence of these estimates on the circadian temperature change, we also investigated average circadian changes of 0.5°F and 1.5°F, instead of 0.9°F. These alternative circadian changes produced morning-vs-evening fever rate ratios in the range of 0.34-0.72, which continue to be consistent with the morning-evening fever rate changes observed in our results, without involving potential biases.

*Summary*

In this appendix, the potential consequences of Berkson's bias and residual confounding were examined through sensitivity analyses, analyses of a general population dataset, and consideration of previous research. Overall, the results are consistent with large morning-evening increases in fever rates that are physiological and not artifacts of Berkson's bias or residual confounding.

### Appendix, COVID-19 Fevers

This appendix provides an overview of fevers in COVID-19, including sections on the occurrence of fever, thresholds used for fever, and time-of-day variation in fever. The included summaries of the literature were last updated on September 21, 2020.

*Occurrence of fever in COVID-19*

Fever is thought to be the most common COVID-19 symptom,[15] and first symptoms often include fever.[16,17] Currently, the amount of evidence on COVID-19 fever rates differs substantially by patient group.

The most evidence on COVID-19 fever rates is available for hospitalized patients, who generally have high rates of fever: 88.7%, including 43.8% on admission;[15] 94.3%, including 87.1% at illness onset;[18] 98.6% at onset;[19] 30.7% on triage or admission;[20] 83%, including 26% on admission;[21] about 89% of symptomatic adult cases;[22] 80.4% of severe and 82.4% of non-severe/common cases at onset;[23] and 85.0% with fever or chills on admission.[24] Fevers in hospitalized patients also present on many days (median fever days per patient: 9 in inpatients without ICU stays,[18] 31 in inpatients with ICU stays,[18] and 12 in surviving inpatients[25]), which would allow multiple opportunities for screening detection if observations are similar outside the hospital setting (which is unknown). Examining the temperatures attained by febrile patients hospitalized with COVID-19[15] suggests that overall they are not unusually high, and do not stand out relative to the temperatures attained in common diseases like seasonal influenza.

Less evidence is available for residents of skilled nursing and assisted living facilities. In studies of skilled nursing facilities, fevers were noted in 39% of residents with confirmed COVID-19,[26] 43% of residents with confirmed COVID-19,[27] and 53% of those under investigation for COVID-19.[27] The lower fever rates may result from older individuals' diminished ability to mount febrile responses.[28]

Limited evidence is also available for the general population of patients with COVID-19 who have not been hospitalized and are not nursing facility residents. However, studies that have tracked new cases suggest fairly high rates of fever in this group, including reports of fever in 71% of contact-traced cases;[29] 75.0% of healthcare personnel, including 41.7% at first onset;[17] 55.4% of healthcare workers according to self-report and 85.0% of healthcare workers defined by temperature ≥37.5°C;[30] at onset, 53.3% of index cases and 56.3% of household members they infected;[31] self-reported by 57.5% of patients at times of positive COVID-19 tests;[32] and self-reported by 48.7% of healthcare workers and 43.7% of others at times of positive COVID-19 tests.[33] Additionally, in CDC analyses, fever was reported for 68% of healthcare personnel with COVID-19[34] and about 73% of symptomatic non-hospitalized adults.[22]

Overall, reports to date show fever rates that are high during COVID-19's clinical course and intermediate at first onset. Our results on the daily cycle of fever rates suggest that some onset research could underestimate fever rates by using morning temperatures, but we cannot tell which studies are affected because none report temperature measurement times.

An added complication is that, in populations with sufficiently low COVID-19 prevalence,* the number of false-positive test results can approach or exceed the number of true-positive test results, leaving open the possibility that false-positive cases may be confused for asymptomatic cases.[35] If this is occurring, it would artificially increase the proportions of COVID-19 cases believed to be asymptomatic (and afebrile) in data from low-prevalence settings. On the other hand, asymptomatic cases are especially likely to go untested in many contexts, creating a contravening bias that could lead to underestimation of the proportion of COVID-19 cases that are afebrile. The competing biases make the overall fever rate in COVID-19 somewhat unclear, despite substantial study.

*Temperature thresholds used for COVID-19*

Several temperature thresholds have been used to define fever for COVID-19, such as ≥100.4°F (≥38.0°C), ≥100.0°F (≥37.8°C), and ≥99.5°F (≥37.5°C). The large range of fever rates reported by previous studies of COVID-19 may be partially attributable to the choice of different fever thresholds in different studies. However, an added complication is that different thermometer sites (e.g., oral, temporal, tympanic, axillary, or rectal) have also been used by different studies, and that some of these sites generally attain lower temperatures in fever than do others.[36] In particular, several studies using low fever thresholds have used axillary thermometers,[25] which tend to show the lowest temperatures during fever. This complication leaves the overall relationship between temperature thresholds and COVID-19 fever rates unclear.

* Or very high rates of testing, such as the frequent retesting that is sometimes given to healthcare workers.

No information is currently available on an optimal fever threshold for COVID-19, overall or by thermometer site.

*Time-of-day variation of fever in COVID-19*

To our knowledge, no study has examined the time-of-day variation of fever in COVID-19. Further, we were unable to find any study of the time-of-day variation of fever in the related disease Severe Acute Respiratory Syndrome (SARS). However, in most febrile diseases, body temperature follows an exaggerated version of the healthy circadian rhythm,[11–13] which reaches its minimum in the morning and its maximum in the late afternoon or evening. (In our study, this pattern of fever rates occurred during both the periods of high influenza activity and the remaining periods.) It remains to be seen whether body temperatures follow this usual pattern in COVID-19, or whether COVID-19 is an exceptional case.

If COVID-19 is an exceptional case, it is possible that its temperature low point may not occur during mornings. In this case, the solutions outlined in our main text may not be useful for COVID-19 prevention, though twice-daily screening could still mitigate the detection problems that are posed by time-of-day variation in fever rates, so long as at least one screening does not occur at the time of temperature low points.

*Recommendations for future research*

Overall, we hope that our research encourages study of fever's course in COVID-19, which could help improve screening practices, such as by identifying optimum times of day for screening. For reasons of practicality, an advantageous time for morning measurements may be directly before leaving for work or school. Based on the physiology of the circadian cycle, an advantageous time for evening temperature measurements could be directly before dinner, which is both near the circadian highpoint and precedes the small metabolism-associated increases in body temperature that follow dinner (and do not result from fever). Before-dinner measurement is also generally consistent with the fever rate highpoints in our analyses of general and influenza-caused fevers.

When planning future COVID-19 fever research, we recommend that investigators keep the following potential obstacles in mind: (1) Inpatients with COVID-19 may not show a daily cycle of fever rates, or may show a distorted cycle. This is because circadian rhythms can be severely disrupted by poor sleep and other stressors of being an inpatient.[37,38] Similarly, night workers may show a distorted or reversed cycle. (2) Observed fever rates can be affected by Berkson's biases (**Appendix, Berskson's bias**). (3) In populations with low COVID-19 prevalence or very high testing rates, many or most of the apparently asymptomatic and afebrile cases may be false positives, as discussed above. (4) Lowering the fever threshold in the morning may help to compensate for the daily cycle of fever rates, but could adversely affect screenings for individuals whose circadian rhythms do not follow the usual pattern of a morning low and evening high, such as some inpatients and night workers.

### Appendix, Screening and Transmission

This appendix uses simple examples to explain how fever and other symptom screenings can confer benefits if they can modestly reduce disease transmission rates during outbreaks, following similar arguments that have been applied for public use of face masks.[39,40]

Temperature screenings are used for COVID-19 because measurements are simple enough to be performed by non-clinicians, because fever is among the most common and earliest symptoms,[15,16] because many or most symptomatic people appear not to self isolate until they receive positive test results,[41,42] because symptomatic health care personnel have often kept working,[17] and because false-negative test results are common,[43] potentially resulting in individuals with symptomatic COVID-19 who continue to participate in work and daily activities because they think they do not have COVID-19. Temperature screenings have also been considered for future pandemic influenza events because fever is a common and early symptom of influenza.[44] However, an important limitation to fever and other symptom screenings is that they cannot detect nonfebrile, asymptomatic, or presymptomatic cases, which are thought to occur substantially for both COVID-19[45] and influenza.[46,47]

Despite symptom screenings' inability to detect some cases, they can still confer benefits that grow considerably in time if they are able to reduce disease transmission rates. For example, suppose that screening only modestly improves case detection and isolation, resulting in an average reduction of 15% in the number of people that each infected individual transmits their disease to.[†] Then, on average, an infected individual would infect 15% fewer others, the people who are nevertheless infected by this individual would infect 15% fewer others, the people who are nevertheless infected by *these* individuals would infect 15% fewer others, and so on, with the benefits of screening compounding at each generation of disease transmission. The consequence is that at the first, second, third, and fourth generations of transmission in a new outbreak, there would be roughly 85%, 72.3%, 61.4%, and 52.2% as many new cases as would otherwise occur (=85%$^n$). In this way, over the span of only a handful of disease transmission generations, the modest 15% reduction in disease transmission translates into the large benefit of having only 52.2% as many new cases as would occur without screening.

The growth of benefits is also why addressing screening failure points, like low morning fever rates, can be more beneficial than intuition may suggest: For example, if addressing this issue were to change the effect on transmission from a 15% reduction to a 25% reduction, then at the fourth generation of a new outbreak, there would be roughly 31.6% as many new case as would occur without screening, rather than 52.2%.

Importantly, the growth of benefits eventually stops following this exponential pattern as the growth of the outbreak stops being exponential itself. However, the benefits slow outbreaks, allowing more time to try case tracking and other limited countermeasures before closures and lockdowns become the only options for stopping extensive disease spread.

---

[†] In other words, this is a reduction of 15% in the disease's *R* effective.

In previous studies, similar reasoning has been pursued with greater thoroughness to explain how large benefits can accompany other imperfect, partial measures of blocking disease transmission, in particular including public use of cloth and procedure face masks to reduce the spread of COVID-19.[39,40] However, we caution that no similar analyses of workplace and school fever screening have been published to date.

### Appendix Methods

This appendix describes additional methods.

*Datasets and measurements*

We performed the Boston study at the Beth Israel Deaconess Medical Center Emergency Department (September 2009–March 2012) using temporal artery thermometers connected to automatic data-loggers that recorded the measured temperatures and times.[3,48] The national study was performed by the US Centers for Disease Control and Prevention as the emergency department components of the year 2003–2010 National Hospital Ambulatory Medical Care Surveys (NHAMCS), which are multi-stage probability sample surveys that provide nationally representative data on hospital emergency and outpatient visits, including visit records from December 2002 to December 2010.[49] NHAMCS collected case records for every *n*th visit following a random start, with the mode of thermometry left to the discretion of participating clinicians and institutions.

For analyses of the Boston study, sample sizes were determined by the study duration. For analyses of the national study, sample sizes were determined by the years chosen for investigation. Years were selected to include a long period during which the survey design and the recorded variables of interest remained consistent enough to analyze together. Years were also chosen to provide some results predating the widespread use of temporal artery thermometers, to demonstrate that time-of-day variations in fever rates were also present beforehand (see original publication[3]).

Please see the original publication[3] that this research letter builds on for further details on patient demographics, dataset characteristics, measurement methods, and inclusion and exclusion choices. The original publication also includes sensitivity analyses that demonstrate robustness of study findings to changes in the exclusion criteria, and to the use of arrival times as substitutes for measurement times in the national study.

*Statistical analyses of the national study*

We accounted for the national study's multistage design to obtain nationally representative findings.[50] For the national study, time-of-day case mix differences in age (years, analyzed with spline), urgency/immediacy of case (4 levels and unknown), pain (4 levels and unknown), sex (male or female), race (black, white, or other), Hispanic or Latino ancestry (yes or no), hospital admission (yes or no), test ordering (yes, no, or unknown), procedure administration (yes, no, or unknown), medication ordering (yes, no, or unknown), ambulance arrival (yes, no, or unknown), and expected payment source (7 categories and unknown) were excluded as responsible factors for the time-of-day fever rate differences using multivariable logistic regression and average marginal predictions.[5,49] Additional variables and categories for gender, race, and ethnicity were not available for some or all study years, and were therefore not analyzed. Additionally, for variables such as urgency/immediacy to be seen and degree of pain, some levels with more detail were merged to obtain consistent categories across study years.

As discussed in the original study,[3] time-of-day variation in the studied characteristics was modest, which helps to explain why the adjusted and unadjusted results were broadly similar.

Cardiovascular events are known to vary by time of day, generally peaking in morning hours.[51] However, the incidence of cardiovascular events was too small to meaningfully affect the time-of-day variation in fever rates (for example, cardiac arrest accounts for 0.17% of emergency department presentations[51]). We therefore decided it was not necessary to control for cardiovascular events in the multivariate analyses.

Anonymity requirements prevented linkage of the Boston temperatures to patient characteristics, thereby preventing multivariate-adjusted analyses of the Boston data.[3,48]

*Statistical analyses with time of day as a continuous variable*

In some appendix figures, time of day was evaluated as a continuous variable instead of being binned (**Supplementary Figures S1, S2, A**). These analyses were performed using logistic regressions with cyclic cubic splines. They account for the national study's multistage design,[50] though national representativeness may not be present in some results from the year-stratified analyses (**Supplementary Figure S2**) because some year-by-outbreak period strata include case records sampled from relatively few hospitals.

Evaluating time as a continuous variable helps to address the arbitrariness of choosing bin boundaries in binned analyses, as well as the possible sensitivity of results to choices of bin boundaries. However, spline results can still be somewhat sensitive to the choices of spline types and parameters, such as knot locations. Additionally, we caution that it is difficult to make inferences about the exact times of the daily minimum and maximum fever rates from the spline fits, owing to statistical uncertainties in these quantities and their potential sensitivity to choices of spline types and parameters, even in large datasets such as used for this study.
